# Transmissibility of a new *Plasmodium falciparum* 3D7 bank for use in malaria volunteer infection studies evaluating transmission blocking interventions

**DOI:** 10.1101/2024.12.18.24319134

**Authors:** Sean A. Lynch, Azrin N. Abd-Rahman, Jenny M. Peters, Juanita M. Heunis, Jeremy Gower, Adam J. Potter, Rebecca Webster, Helen Jennings, Susan Mathison, Nischal Sahai, Fiona H. Amante, Bridget E. Barber

**Affiliations:** QIMR Berghofer Medical Research Institute, Brisbane, Queensland, Australia; Queensland University of Technology (QUT), Faculty of Health, School of Biomedical Sciences, Brisbane, Queensland, Australia; University of the Sunshine Coast Clinical Trials Centre, Brisbane, Queensland, Australia

**Keywords:** malaria transmission, gametocyte, anopheles, oocysts, controlled human malaria infection, clinical trial

## Abstract

Transmission blocking activity is an important characteristic of antimalarial drugs, and can be evaluated in malaria volunteer infection studies (VIS). We undertook a pilot VIS to evaluate the suitability of a recently manufactured *P. falciparum* 3D7 bank (3D7-MBE-008) for evaluating transmission blocking interventions. Four adults were inoculated with *P. falciparum* 3D7-MBE-008 infected erythrocytes and administered piperaquine on days 8 and 10 to clear asexual parasitemia while allowing gametocyte development. On day 25, participants were randomised (1:1) to receive either 0.25 mg/kg primaquine (primaquine group) or no intervention (control group). Transmissibility was assessed by enriched membrane feeding assays on days 25, 29, 32, and 39, with transmission intensity (proportion of mosquitoes infected) determined by 18S qPCR. All participants were infective on day 25, with a median 94% (range, 12%–100%) of mosquitoes positive for oocysts, and 76% (range, 8%–94%) positive for sporozoites. In the primaquine group, mosquito infectivity decreased substantially between days 25 and 29. In the control group, mosquito infectivity remained high up to day 32, and persisted to day 39 in one participant. The *P. falciparum* 3D7-MBE-008 parasite bank induced blood-stage infections that were highly transmissible to mosquitoes and is therefore suitable for evaluating transmission blocking interventions.

*Trial registration* anzctr.org.au (registration number: ACTRN12622001097730), registered 08/08/2022.

## INTRODUCTION

Despite continued advances in the fight against malaria, the global burden of the disease remains high, with an estimated 249 million cases and 608,000 deaths in 2022 [1]. The spread of antimalarial resistance and mosquito vector species, in addition to the impact of climate change, represent major threats to malaria control [1]. New interventions are urgently required to accelerate progress toward malaria elimination, including strategies that can disrupt transmission of parasites from humans to mosquitoes [2].

Fortunately, a number of promising new antimalarial agents are progressing through the development pipeline [3], and the ability to test the transmission blocking activity of these novel agents will be essential for informing partner drug selection and optimal use of these agents. The transmission blocking activity of antimalarial agents has been assessed in Phase 2/3 studies, generating direct evidence of effectiveness against naturally acquired infections in endemic settings [4, 5, 6, 7, 8]. However, this approach is associated with significant challenges, including the confounding effects of adjunctive curative therapy, pre-existing immunity in participants [9], variation in mosquito infectivity [10], as well as logistical, infrastructure and regulatory considerations [11].

Malaria volunteer infection studies (VIS) enable clinical assessment of interventions in malaria-naïve participants in a well-controlled environment [12]. Although VIS have predominantly focused on assessing the activity of antimalarials against blood-stage asexual parasites [13, 14, 15, 16], the utility of VIS for evaluating transmission blocking interventions has recently been demonstrated [17, 18], with blood-stage VIS shown to be more suitable than sporozoite VIS due to higher gametocyte densities obtained in the former [19]. The majority of blood-stage malaria VIS have been conducted using the *P. falciparum* 3D7-V2 parasite bank, produced in 1994 with donated blood from an infected volunteer [20]. However, due to limited supply of this bank, a new bank (3D7-MBE-008) was manufactured using a wave bioreactor to expand the 3D7-V2 parasites in vitro. Although a pilot study confirmed the infectivity of the new 3D7-MBE-008 bank [21], the transmissibility to mosquitoes was not assessed.

Long-term in vitro cultivation of *P. falciparum* can lead to mutations that impair production and development of gametocytes [22, 23] and transmissibility to mosquitoes. Furthermore, gametocytes cultured in vitro differ markedly in their gene expression to those of the same parasite line during naturally acquired infection [24, 25, 26]. Thus, the ability of a parasite line to develop gametocytes (and transmit to mosquitoes) in a blood-stage infection may not be ascertainable by observations of in vitro culture. We therefore undertook a pilot malaria VIS to evaluate the transmissibility of the 3D7-MBE-008 parasite bank and its suitability for use in future studies evaluating transmission blocking interventions.

## METHODS

### Study design and participants

The study design is shown in Fig. 1. This was a randomized, open-label clinical trial that utilised the induced blood stage malaria (IBSM) model to evaluate the transmissibility of *P. falciparum* 3D7-MBE-008 parasites from participants to mosquitoes. Healthy malaria-naïve males and females, aged 18–55 years, and with >30% of normal range G6PD enzyme activity levels, were eligible for inclusion. The study was conducted in accordance with relevant guidelines and regulations, including the Declaration of Helsinki (as revised in 2013), Australian National Health and Medical Research Council (NHMRC) National Statement on Ethical Conduct in Human Research (as revised in 2018) and the Integrated Addendum to ICH E6(R1): Guideline for Good Clinical Practice ICH E6(R2) (2016). The study was conducted at the University of Sunshine Coast Clinical Trials Centre (Brisbane, Australia) and was approved by the Human Research Ethics Committees of QIMR Berghofer Medical Research Institute (Reference Number P3821), University of Sunshine Coast (Approval Number A221785) and Lifeblood (Reference Number 2022#13). The study was registered on the Australian New Zealand Clinical Trials Registry (ACTRN12622001097730). All participants gave written informed consent.

**Figure 1.**
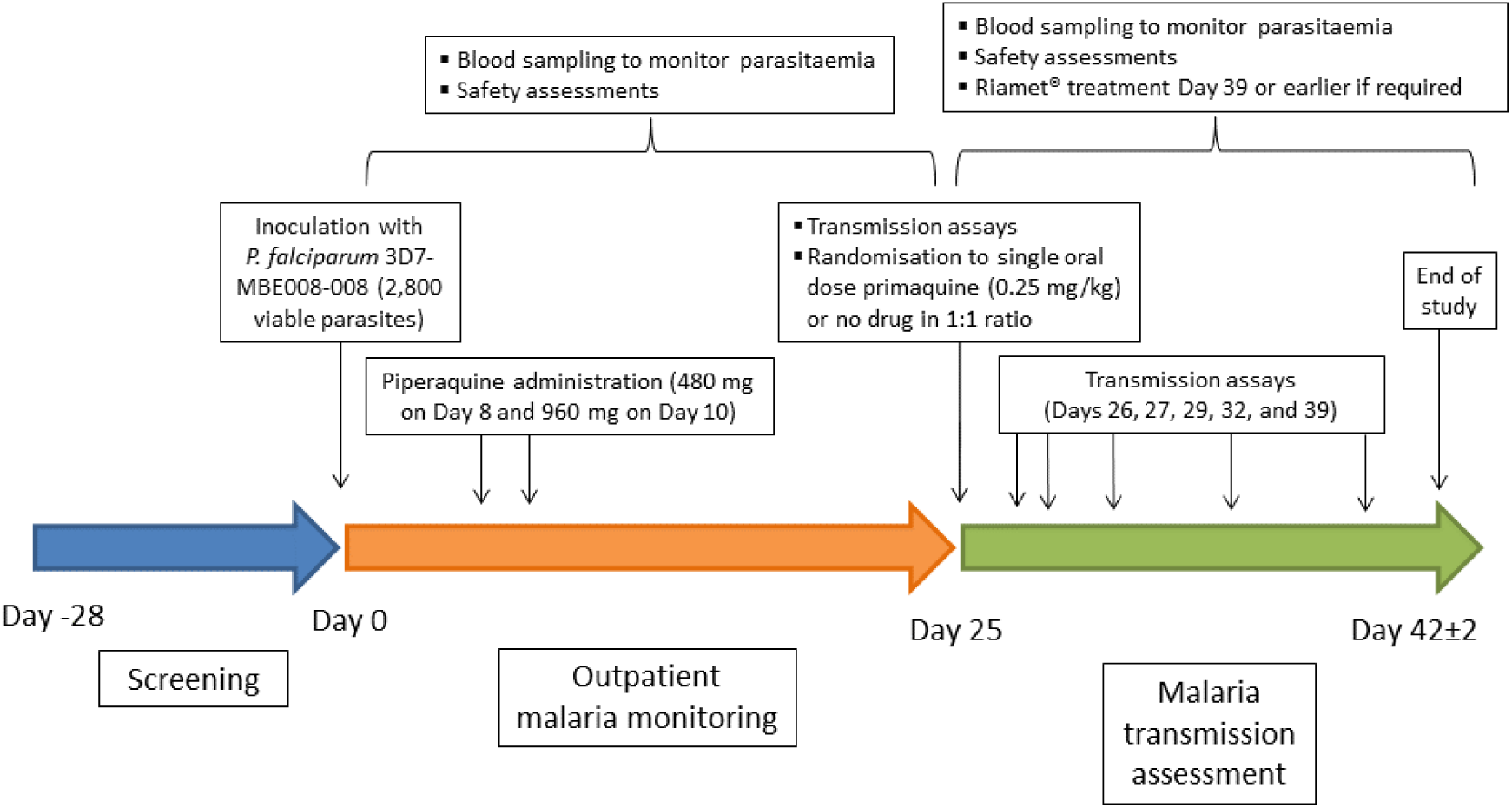
Schematic representation of the study design.

### Procedures

Participants were intravenously inoculated with approximately 2,800 viable *P. falciparum* 3D7-MBE-008 infected erythrocytes on day 0. Parasitemia was monitored daily by qPCR targeting the 18S rRNA gene of *P. falciparum* [27] from day 4. Piperaquine phosphate tablets (Copea Pharma) were administered on days 8 (480 mg) and 10 (960 mg) to clear asexual parasitemia while allowing development of mature gametocytes [17, 28]. From days 11 to 24 parasitemia was monitored 3 times per week, including determination of parasite lifecycle stage by qRT-PCR targeting the female gametocyte-specific transcript *pfs25* and male gametocyte-specific transcript *pfMGET* [29], and the ring stage specific transcript *pfSBP1* [30]. On day 25, transmission of gametocytes to mosquitoes was assessed by enriched membrane feeding assay (eMFA). Participants were then randomised in a 1:1 ratio (randomisation list generated electronically by the study statistician) to receive either a single oral dose of approximately 0.25 mg/kg primaquine (Primacin, Boucher & Muir) to clear gametocytes [17] (primaquine group), or no transmission blocking intervention (control group). Additional eMFAs were performed on days 26, 27, 29, 32, and 39; however, data from days 26 and 27 have been excluded due to the malfunction of a critical piece of equipment on these days resulting in the gametocyte samples exceeding the maximum temperature for viability.

Compulsory rescue treatment with artemether-lumefantrine (Riamet, Novartis Pharmaceuticals) was administered to all participants following the final eMFA on day 39, or earlier in the event of recrudescence of asexual parasites. Participants were administered 45 mg primaquine at the end of study (day 42) if required to clear residual gametocytes.

### Infection of *Anopheles stephensi* mosquitoes

Transmission of gametocytes to *An. stephensi* mosquitoes was evaluated by eMFA following collection of 60 mL of venous blood from participants at designated time points. Blood was leucodepleted by Plasmodipur filtration (EuroProxima), adjusted to 50% hematocrit with RPMI-1640 and layered over a 65% Percoll density gradient followed by spinning at 1500 × *g* for 15 min at 37°C. The resulting enriched gametocytes were then washed with RPMI-1640 and reconstituted in malaria naïve donor erythrocytes and serum to a volume of 1.3 mL at 50% hematocrit to form the mosquito feed mix [18].

Female *An. stephensi* mosquitoes 4–5 days post-emergence were distributed into pots and provided an 8% sucrose solution containing 0.1% methylparaben (Sigma) for 24 hours, followed by 5–6 hours starvation prior to membrane feeding. Two separate pots containing approximately 75 mosquitoes were fed on each enriched participant feed mix sample. Mosquitoes were fed for 30 minutes through stretched Parafilm membranes on water jacketed glass feeders attached to a heated circulating water bath (36–37°C). Blood fed mosquitoes were subsequently provided an 8% sucrose/0.05% para-aminobenzoic acid (Sigma) solution and maintained at approximately 27°C and 75% relative humidity with a 12:12 hour light-dark cycle. Fifty mosquitoes from one pot per participant were collected on day 8 post-feed to assess oocyst infection by 18S qPCR of whole mosquito samples. The presence of oocysts in the remaining mosquitoes was visually confirmed by microscopic examination of dissected midguts stained with 0.5% mercurochrome solution (Supplementary Fig. S1). From the second pot, 50 mosquitoes were dissected after 16 or 17 days to isolate head-thorax samples for 18S qPCR. The presence of sporozoites in the remaining mosquitoes was confirmed by microscopic examination of isolated salivary glands (Supplementary Fig. S1). All mosquitoes were visually examined to confirm gravidity prior to analysis ensuring that only blood-fed mosquitoes were analysed.

### DNA extraction and qPCR analysis of mosquito tissue

DNA from whole mosquitoes (oocyst stage analysis) or dissected head/thorax samples (sporozoite analysis) was extracted using the Maxwell® RSC PureFood GMO and Authentication Kit (Promega). Samples were collected into 195 μL CTAB buffer and 5 μL DNA Extraction Control (DEC) (Meridian Bioscience) was added prior to processing. Samples were homogenised with a Mini Beadbeater (Sigma Aldrich) for 90 seconds using 2.3 mm zirconia/silica beads. 200 μL CTAB buffer, 5 μL RNAse A, and 20 μL Proteinase K were added and samples incubated for 30 minutes at 65°C. DNA was extracted as per Manufacturer’s instructions using the Maxwell® RSC Instrument (Promega) and eluted into 100 μL elution buffer. qPCR assays were performed as previously described [18].

### Outcomes

The primary outcome was the proportion of participants who transmitted to mosquitoes on day 25. Secondary outcomes included the intensity of mosquito infection per participant on day 25; the within-person percentage change in mosquito infection intensity on day 26, 27, 29, 32 and 39, compared to day 25; the within-person percentage change in gametocyte density on day 26, 27, 29, 32 and 39, compared to day 25; and the incidence and severity of adverse events.

### Statistical analysis

The small sample size of this pilot study was chosen based on the primary objective of assessing the transmissibility of *P. falciparum* 3D7-MBE-008 from humans to mosquitoes. Four participants were considered sufficient to meet this objective, based on a previous study evaluating the transmissibility of the *P. falciparum* 3D7-V2 malaria cell bank (MCB) from which the 3D7-MBE-008 MCB was produced, demonstrating high intensity transmission to mosquitoes in 6/6 participants [18].

The intensity of mosquito infection was calculated as the proportion of infected mosquitoes per participant. For each participant, the within-person change in intensity of mosquito infection at each time point, compared to baseline (day 25), was reported as a percentage. Gametocyte density was reported as transcripts per mL or gametocytes per mL (by 18S qPCR) and the within-person reduction from baseline reported as a percentage. Spearman’s correlation was used to assess whether gametocyte density was associated with transmission intensity on day 25. The relationship between oocyst and sporozoite infectivity was assessed using repeated measures correlation following logit(x+0.01) transformation, to account for multiple measures for each participant [31]. Statistical analyses were conducted using R statistical package version 4.3.1 (R Core Team, 2024).

### Post-hoc analysis

Due to the recrudescence of asexual parasitemia that occurred in 2 participants, a population pharmacokinetic (PK) model of piperaquine [32] was used to devise a piperaquine dosing regimen for future studies that would maintain piperaquine plasma concentrations above minimum inhibitory concentration (MIC) of 4.9 ng/mL [32] and hence prevent parasite recrudescence throughout the transmission period (up to day 32 post-inoculation). The piperaquine dosing regimen used in the current study (480 mg on day 8, 960 mg on day 10) and additional doses of 480, 640 and 960 mg given at days 11–17 were simulated in 500 hypothetical participants with body weight of 74 kg (median body weight in previous malaria VIS) over 40 days. Individual PK parameters for the hypothetical malaria VIS participants were sampled from uncertainty and variability distributions and were used to simulate individual PK profiles according to each dosing regimen. PK profiles were plotted and time above MIC (4.9 ng/mL) was determined for various dosing regimens.

## RESULTS

### Participants

The study was conducted from October to December 2022. Eleven volunteers were screened (Supplementary Fig. S2 and Supplementary Table S1). Five volunteers did not meet the eligibility criteria; 2 met the eligibility criteria and acted as reserves on the day of malaria inoculation. Four participants (3 males, 1 female; aged 31 to 45 years) were enrolled, completed the study, and were included in analysis of study endpoints.

### Parasitemia

Parasites were first detected 4–5 days after inoculation in all participants (Fig. 2a-d), and parasite density increased up to a median of 38,692 parasites/mL (range, 8,096–743,372 parasites/mL) on day 8 when all participants were administered 480 mg piperaquine. As expected, this initial dose did not resolve asexual parasitemia and a second larger dose of piperaquine (960 mg) was administered on day 10. Asexual parasitemia was initially cleared in all participants following this second dose; however, parasite recrudescence was observed on days 18 and 29 in Participants 4 and 2, respectively. These participants both received a curative regimen of artemether-lumefantrine on day 32. Female gametocytes were detected on day 10 in Participants 2, 3 and 4, and by day 13 all participants had circulating female and male gametocytes (Fig. 2e-h). All participants were aparasitemic by 18S qPCR at the end of study.

**Figure 2.**
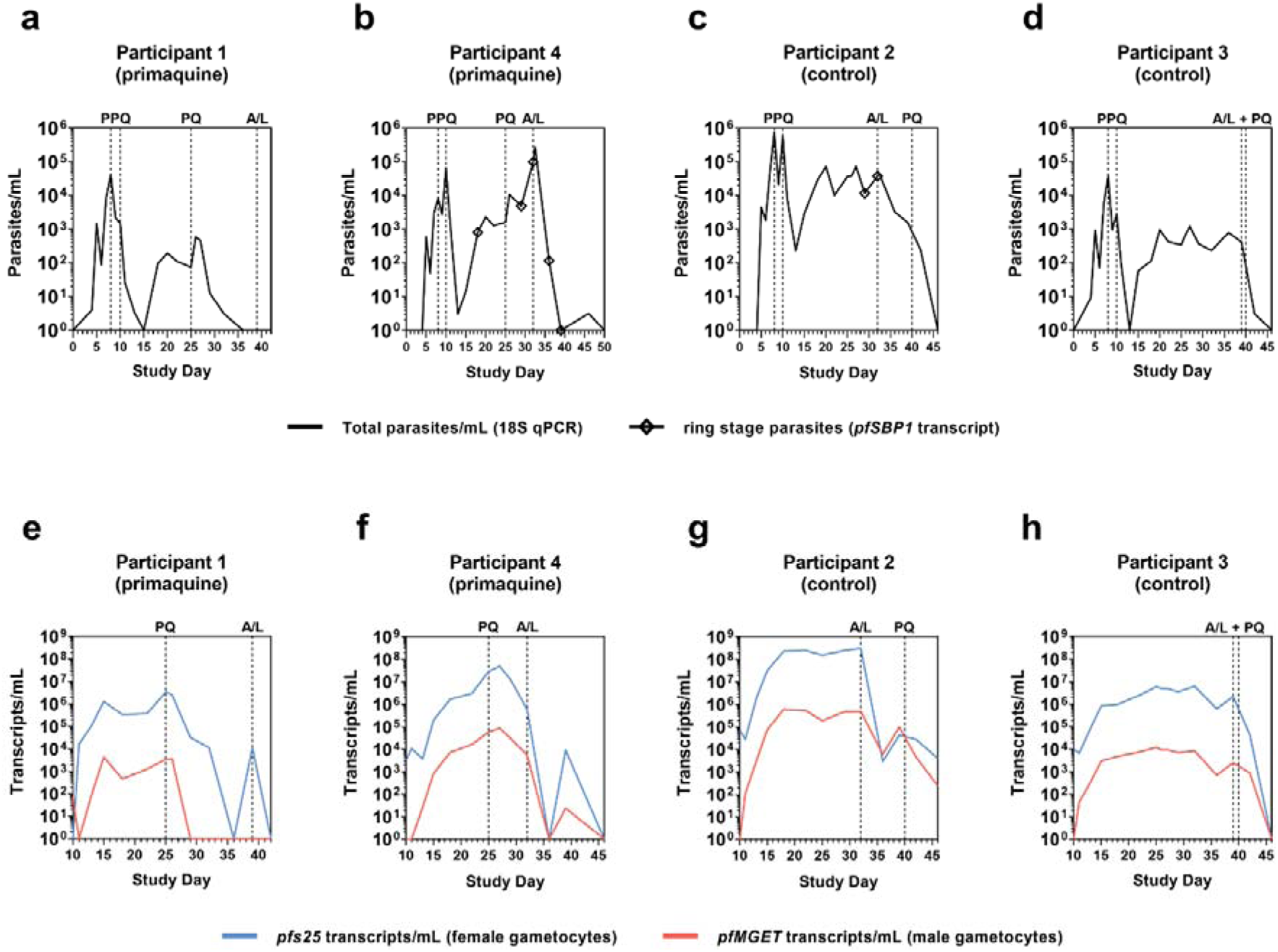
Parasitemia-time profiles by participant. Solid black lines indicate total parasitemia as measured by 18S qPCR (**a**-**d**); diamonds indicate detection of ring-stage parasites by *pfSBP1* (**a**-**d**); solid blue lines indicate female gametocyte transcript *pfs25* and solid red lines indicate male gametocyte transcript *pfMGET* density (**e-h**). Drug administration time points are indicated by dashed vertical lines. Abbreviations: PPQ, piperaquine; A/L, artemether-lumefantrine; PQ, primaquine.

On day 25 Participants 1 and 4 were randomised to receive primaquine (15 mg and 22.5 mg respectively, given body weights of 73.8 kg and 88.2 kg, respectively), while Participants 2 and 3 were randomised to receive no transmission blocking intervention. In the primaquine group, Participant 1 experienced a sharp decline in female and male gametocytes from day 26, with male and female gametocytes being undetectable by day 29 and day 36, respectively (Fig. 2e). Female gametocytes were again detected on day 39, but cleared by day 42 following administration of artemether-lumefantrine. Participant 4 experienced a reduction in female and male gametocytes from day 27 (Fig. 2f). However, the reappearance of asexual parasites on day 29 necessitated early rescue treatment with artemether-lumefantrine on day 32 (Fig. 2b).

In the control group, gametocyte density remained relatively stable in both participants from day 25 up until administration of artemether-lumefantrine (Fig. 2g, h). In Participant 3 this occurred as scheduled on day 39, while in Participant 2 artemether-lumefantrine was administered on day 32 due to parasite recrudescence. Gametocyte densities declined following administration of artemether-lumefantrine, although both participants required administration of primaquine (45 mg) prior to end of study to clear residual gametocytes. The within-person change in gametocyte density (as measured by 18S qPCR) for each participant is presented in Table 1.

**Table 1.**
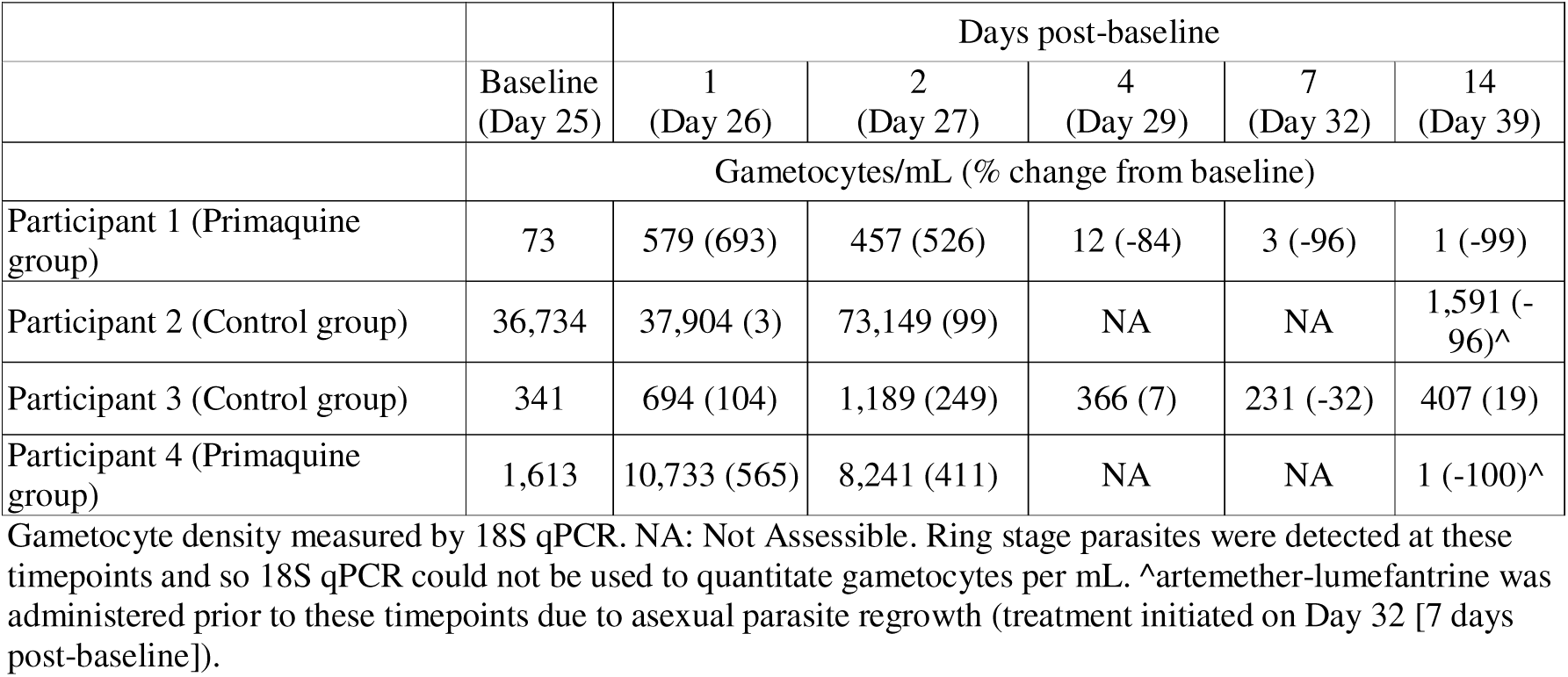
Gametocyte density and change from baseline during the malaria transmission analysis period.

### Mosquito transmission

Gametocytes in participants’ blood samples were successfully enriched using a Percoll density gradient and fed to mosquitoes. All participants were infectious to mosquitoes on day 25 at both the oocyst level and the sporozoite level (Fig. 3). The intensity of mosquito infection was high, with a median 94% (range, 12%–100%) of mosquitoes positive for oocysts, and median 76% (range, 8%–94%) positive for sporozoites. A positive trend was identified between the intensity of mosquito infection and gametocyte density on day 25 at the oocyst level (r=1.0, p=0.083) and the sporozoite level (r=1.0, p=0.083).

**Figure 3.**
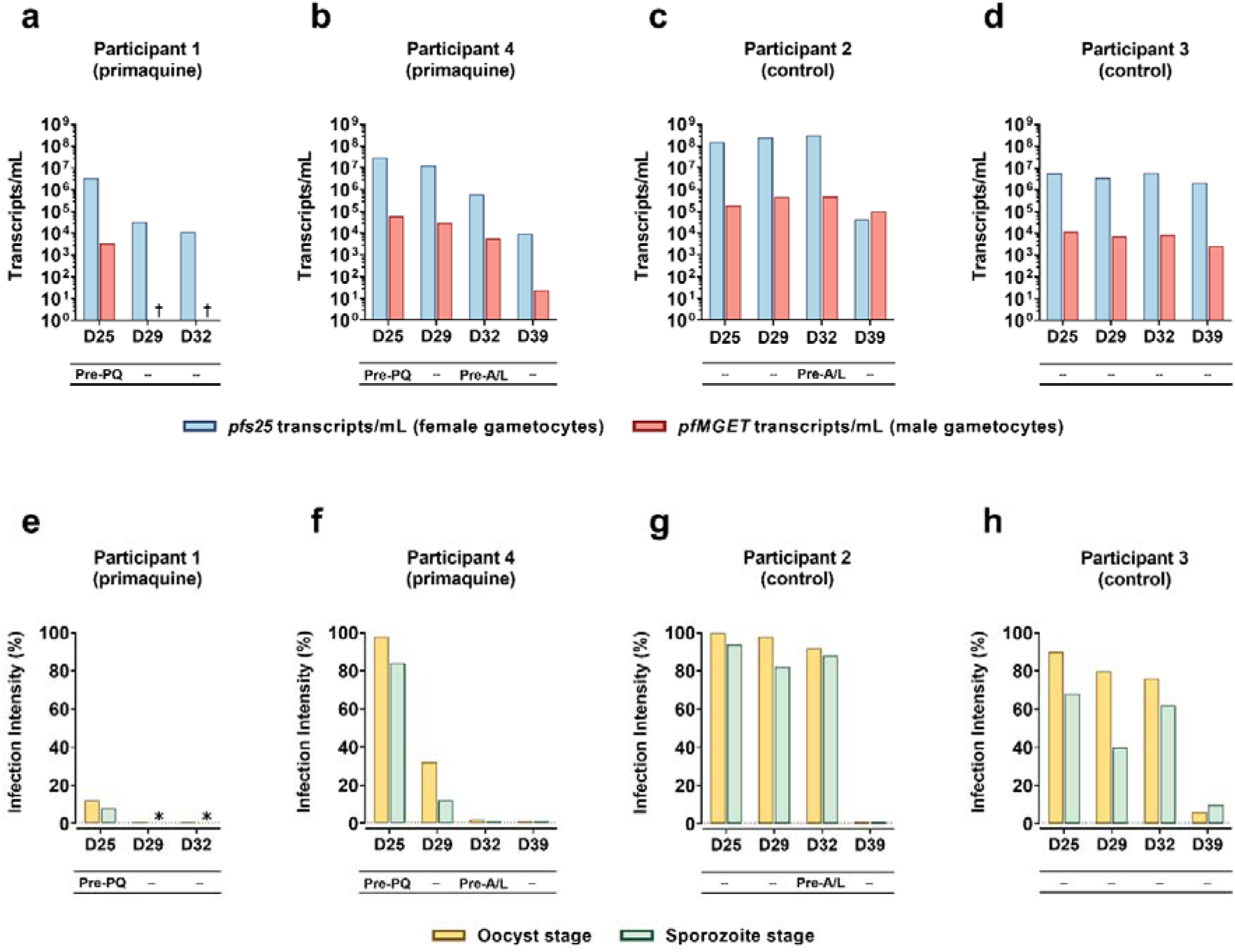
Development of gametocytemia and transmissibility to *An. stephensi* mosquitoes. Female gametocyte specific transcript *pfs25* (blue bars) and male *pfMGET* transcript density (red bars) of each participant at transmission time points (**a-d**). Infection intensity at oocyst stage (yellow bars) and sporozoite stage (green bars) (**e-h**). Baseline transmission assays were conducted on day 25 before Participants 1 and 4 were administered a single dose of 0.25 mg/kg primaquine. Participant 2 and Participant 4 were administered artemether-lumefantrine on day 32 as a rescue treatment following asexual parasite regrowth. Abbreviations: A/L, artemether-lumefantrine; PQ, primaquine. †: no *pfMGET* transcripts were detected at these time points. *: sporozoite stage mosquitoes from these time points were not analysed given that no oocysts were detected.

In the primaquine group, mosquito infectivity decreased substantially between day 25 and day 29 in both participants (Fig. 3e, f). At the oocyst level, infectivity decreased from 12% to 0% in Participant 1 and from 98% to 32% in Participant 4, with a further reduction to 2% by day 32 in Participant 4. At the sporozoite level, infectivity in Participant 4 decreased from 84% to 12% between days 25 and 29, and further to 0% by day 32. Mosquitoes were not examined for sporozoites beyond day 25 in Participant 1 due to the absence of oocysts.

In the control group, intensity of transmission remained high in both participants up to day 32, with 92% and 76% of mosquitoes positive for oocysts in Participants 2 and 3, respectively, and 88% and 62% of mosquitoes positive for sporozoites (Fig. 3g, h). Early rescue treatment with artemether-lumefantrine was administered on day 32 to Participant 2 following asexual parasite recrudescence, and on day 39 no mosquito infection was observed; however, Participant 3 remained infective up to day 39 (31 days after the first dose of piperaquine), with 6% and 10% of mosquitoes positive for oocysts and sporozoites, respectively.

The oocyst and sporozoite infection rates by 18S qPCR were strongly correlated (r_rm_=0.97, [95% confidence interval, 0.88–0.99], p<0.001).

### Modelling piperaquine dosing strategies to avoid parasite recrudescence

Simulations were performed as a post-hoc analysis to predict the time period that piperaquine plasma concentrations would remain above the MIC (4.9 ng/mL) with different dosing regimens (Fig. 4 and Supplementary Table S2). The piperaquine dosing regimen used in the current study (initial dose of 480 mg and second dose of 960 mg 2 days later) was predicted to have a 95% probability of maintaining plasma concentrations above the MIC for 13.25 days (Supplementary Table S2). The optimum dosing regimen was determined to be an additional dose of 960 mg administered at 9 days after the initial 480 mg dose, with this dosing regimen predicted to have a 95% probability of piperaquine concentrations above the MIC for 24 days (equivalent to day 32 in the current study) (Fig. 4).

**Figure 4.**
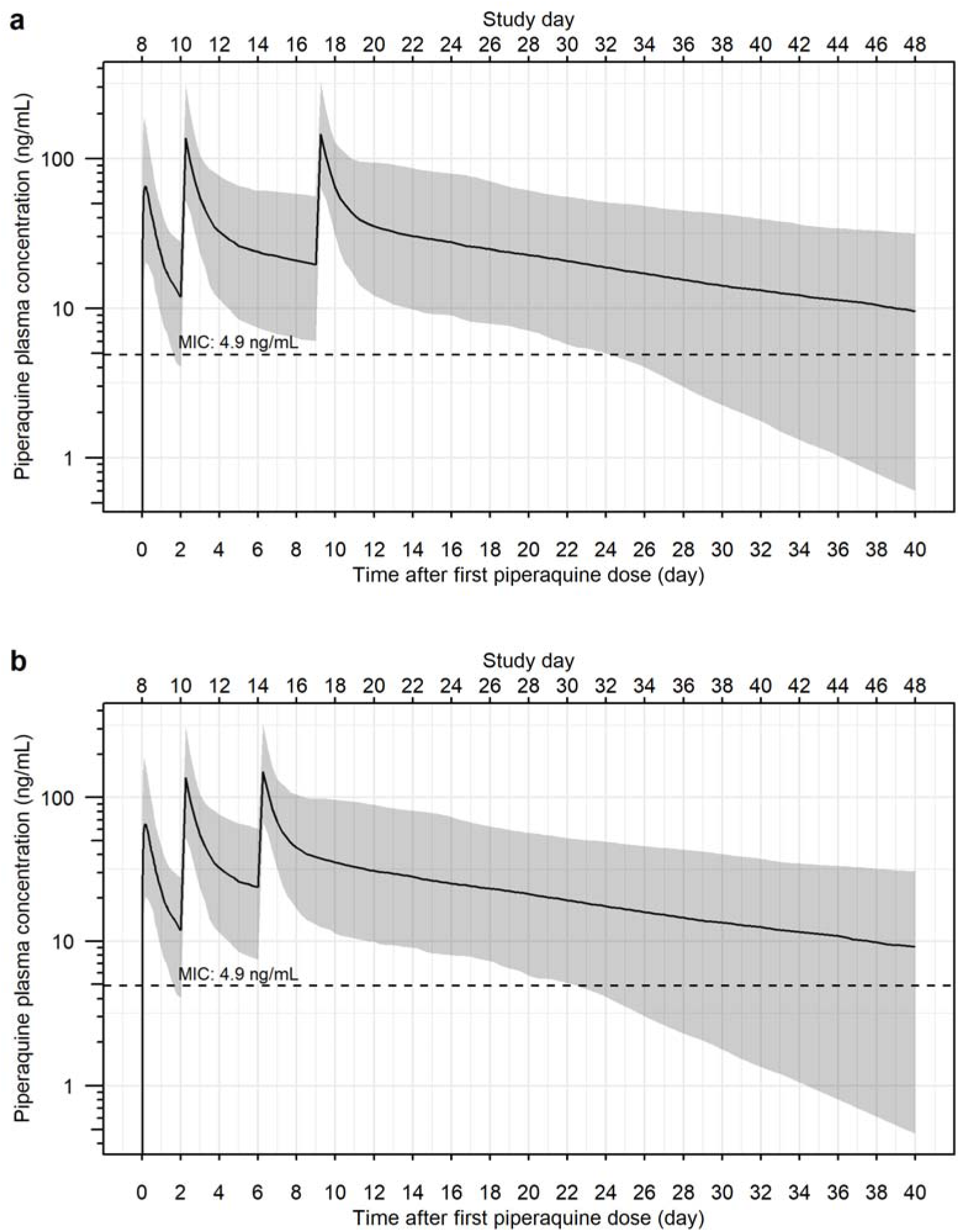
Simulated piperaquine plasma concentration-time profiles relative to study day (upper horizontal axes) and time after first dose (lower horizontal axes). An additional dose of 960 mg administered 9 days (**a**) post-initial dosing was predicted to have ≥95% probability of sufficient piperaquine exposure for up to 32 days post-inoculation, thereby preventing asexual parasitemia recrudescence during mosquito transmission assessment. The piperaquine plasma concentration time profile for administration of 960 mg of piperaquine 6 days post initial dosing is also shown (**b**). The solid line and the shaded area represent the median concentration and 95% prediction interval, respectively. The dashed line represents the piperaquine minimum inhibitory concentration (MIC) [32].

### Safety and Tolerability

All participants experienced at least one adverse event (AE) (Supplementary Table S3). Of the 53 AEs reported, most were mild or moderate (51/53) and considered related to malaria (38/53). There were no AEs or serious adverse events leading to the discontinuation of the trial. Two participants experienced a severe (grade 3) AE, with both of these being a decrease in lymphocyte count. Both AEs were considered to be related to the malaria challenge agent and spontaneously resolved within 7 days.

## DISCUSSION

This study evaluated the transmissibility of a new *P. falciparum* 3D7 blood-stage parasite bank (MBE-008) to determine its suitability for evaluating transmission blocking interventions in future malaria volunteer infection studies (VIS). We demonstrated that transmissibility of MBE-008 parasites was high, with gametocytes infective to mosquitoes in all four participants on day 25 following inoculation. Furthermore, intensity of transmission was high, with a median 94% (range, 12%–100%) of mosquitoes positive for oocysts, and 76% (range, 8%–94%) positive for sporozoites. This study has therefore demonstrated that this parasite bank is suitable for evaluating transmission blocking interventions in malaria VIS.

The transmissibility of this new *P. falciparum* 3D7-MBE-008 parasite bank is comparable to that demonstrated for the *P. falciparum* 3D7-V2 bank, which has been utilized for numerous malaria VIS including several studies evaluating transmission blocking interventions [17, 18, 33]. In the most recent transmission VIS utilising the 3D7-V2 bank and conducted using the same study design as the current study, transmission to mosquitoes on day 25 post-inoculation was demonstrated in 6/6 participants, with transmission intensity also being high (median 86% [range, 22%–98%] of mosquitoes positive for oocysts and 57% [range, 4%– 92%] positive for sporozoites) [18]. Taken together, with this study design (albeit with a different *P. falciparum* 3D7 bank), transmission has therefore been demonstrated in 10 of 10 participants, with high intensity transmission (median 89% [interquartile range, 61%–98%] of mosquitoes positive for oocysts and 70% [interquartile range, 32%–91%] positive for sporozoites across these 2 studies). The current study has therefore further confirmed the utility of the malaria VIS for evaluating transmission blocking interventions.

Primaquine is currently the only antimalarial recommended for the purpose of transmission blocking [34]. Therefore, we took the opportunity to observe the effect of primaquine on gametocyte density and transmission to mosquitoes, as well as the duration of gametocytemia and transmission to mosquitoes in the absence of any transmission blocking intervention. Given the small sample size of this pilot study, we did not intend to make definitive comparisons between treatment groups. For the 2 participants administered primaquine (0.25 mg/kg), the decline in gametocytemia was relatively modest, with gametocytes persisting at a high level 4 days post-primaquine in one participant. This delay in clearance of gametocytes is consistent with previous studies in malaria-endemic regions involving supplementation of antimalarial regimens with a single low dose of 0.25 mg/kg primaquine as a transmission blocking agent [4, 5, 7]. In these studies, gametocyte densities reduced significantly between 2–7 days of treatment, but persisted up to 28 [4, 7] or 42 days [5]. Importantly, the continued presence of gametocytes after primaquine treatment has not been shown to correlate with mosquito transmission, suggesting sterilisation of gametocytes preceding their clearance [35, 36].

In both participants administered primaquine, substantial reductions in transmission were observed by day 29 (4 days post-primaquine). These results are generally consistent with two Phase 2 studies in patients in malaria-endemic regions, where the same dose of primaquine administered alongside antimalarial treatment resulted in complete block in transmission within 2 days [5, 7]. The low level of transmission demonstrated 4 days post-primaquine in one participant in the current study likely reflects the fact that transmission intensity in this participant was particularly high at baseline (98% of mosquitoes positive for oocysts), in comparison to the substantially lower baseline transmission intensities reported in the Phase 2 studies (medians of 11%–27% and 23%–24% reported by Dicko et al. [5] and Stone et al. [7], respectively). The high levels of transmission observed in our study were likely aided by our gametocyte enrichment process. In a separate Phase 2 study, transmission intensity between non-enriched and enriched participant samples was compared before and after antimalarial treatment supplemented with low dose primaquine (0.25 mg/kg) [37]. Following treatment, 2 participants that were initially non-infectious were able to infect mosquitoes following gametocyte enrichment. While the high intensity of transmission observed in malaria VIS may result in more prolonged transmission following a transmission blocking intervention than observed in Phase 2 studies, the relative reductions in transmission are comparable. Furthermore, the high intensity of transmission at baseline in malaria VIS increases the power to evaluate transmission blocking interventions in these studies, enabling interventions to be evaluated with relatively low samples sizes.

We also evaluated gametocytemia and transmission to mosquitoes in 2 participants who were not administered any transmission blocking intervention. The duration of transmission in these 2 participants was long, with a high level of transmission demonstrated 22 days following the first dose of piperaquine, and one participant still infectious to mosquitoes 31 days following the first dose of piperaquine. These findings demonstrate the longevity of gametocytes in the absence of gametocytocidal treatment, consistent with the major contribution of asymptomatic parasitemia/gametocytemia to ongoing transmission in malaria-endemic regions [38, 39, 40, 41], and highlight the critical importance of incorporating transmission blocking interventions into malaria control programs.

An important observation in this study was that 2 participants experienced recrudescence of asexual parasites following administration of 2 doses of piperaquine, confounding the analyses of gametocyte dynamics and infectivity to mosquitoes between and within participants. We therefore performed simulations to estimate a piperaquine dosing strategy that would prevent parasite recrudescence in future studies. These simulations determined that a three-dose regimen, with 960 mg administered 2 days and 9 days following the initial 480 mg dose, will have a high likelihood of preventing recurrence of asexual parasitemia for a sufficient duration to assess transmission blocking interventions. This analysis will inform the design of future malaria VIS evaluating transmission blocking interventions.

In addition to the recrudescence of asexual parasitemia that occurred in 2 participants, this study had other limitations. First, transmission was not assessed as planned on days 26 and 27 (1 and 2 days following administration of primaquine), limiting our ability to assess the early transmission blocking activity of primaquine. Second, as stated above, the small sample size of this pilot study meant that there was insufficient statistical power to definitively assess the impact of primaquine on gametocytemia or transmission to mosquitoes.

In summary, this study demonstrated the transmissibility of a new *P. falciparum* 3D7 parasite bank (MBE-008), confirming its suitability for use in future malaria VIS evaluating transmission blocking interventions. Together with another recent study using the same study design with a different *P. falciparum* 3D7 bank, transmission to mosquitoes has now been demonstrated in 10 of 10 participants, with a high level of transmission intensity. Taken together these studies have confirmed the utility of malaria volunteer infection studies for evaluating transmission blocking interventions.

## DATA AVAILABILITY

Data supporting the findings of this study are available in the Supplementary Information files and from the Corresponding Author upon request.

## ACKNOWLEDGMENTS

We thank all the volunteers who participated in the study; staff at University of the Sunshine Coast Clinical Trials Centre who conducted the trial; Ms Hayley Mitchell for assistance with transmission experiments; staff at Queensland Paediatric Infectious Diseases laboratory for qPCR analysis; and Ms Ria Woo for managing clinical data.

## AUTHOR CONTRIBUTIONS

S.A.L., A.J.P., R.W., F.H.A., and B.E.B. designed the study. S.A.L., J.M.P., J.M.H., and J.G. performed experiments. S.A.L. and J.M.P. performed molecular analyses. S.A.L., A.N.A.R., and B.E.B. analysed data. R.W., H.J., S.M., N.S., F.H.A., and B.E.B. managed and coordinated research activities. S.M., N.S., and B.E.B. conducted the clinical trial. S.A.L. wrote the first draft of the manuscript. B.E.B. revised the draft manuscript. All authors reviewed the manuscript prior to publication.

