## Supplementary Information for "Transmissibility of a new *Plasmodium falciparum* 3D7 bank for use in malaria volunteer infection studies evaluating transmission blocking interventions"

**Supplementary Figure S1.** Oocysts and sporozoites visualised by microscopy. **(a)** Representative midgut from sampled mosquitoes fed by enriched membrane feeding assay (eMFA) on Participant 2, Day 25. **(b)** Isolated salivary glands from sampled mosquitoes fed by eMFA on Participant 2, Day 32. Sporozoites were visualised by phase contrast microscopy.

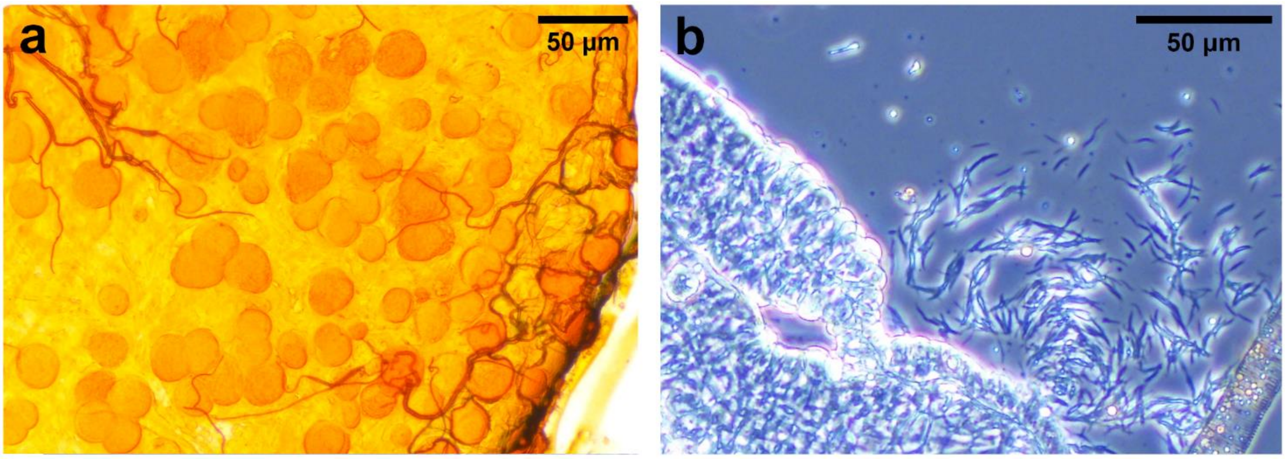

**Supplementary Figure S2.** Study Profile.

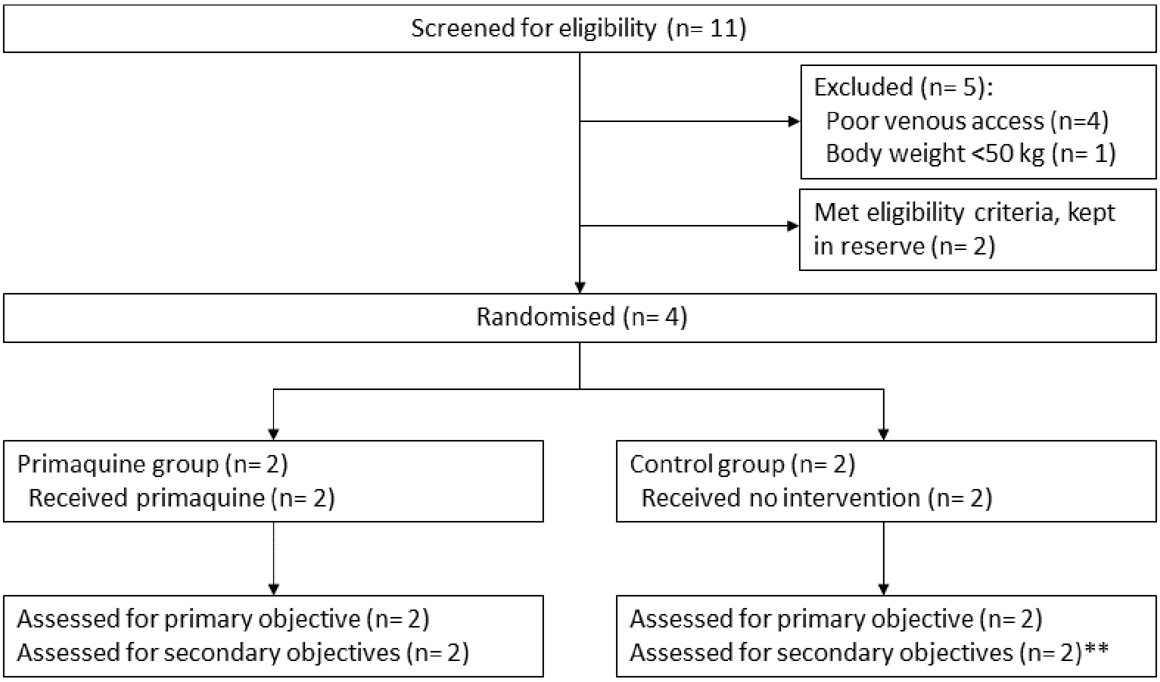


**: The mosquito feeding assay for Participant 1 on Day 39 was not conducted due to absence of parasites detectable by 18S qPCR.

### Supplementary Table S1. Participant Demographic Characteristics

| | **Participant 1** | **Participant 2** | **Participant 3** | **Participant 4** |
|---|---|---|---|---|
| Age (years) | 31-35 | 41-45 | 31-35 | 36-40 |
| Sex | Male | Female | Male | Male |
| Race | White | White | White | White |
| Weight (kg) | 73.8 | 66.9 | 68.9 | 88.2 |
| Body mass index ($kg/m^2$) | 26.2 | 22.9 | 25.3 | 26.9 |

**Supplementary Table S2. Lower Limit of 95% Prediction Interval of Time (in Days) Above Piperaquine Minimal Inhibitory Concentration For Simulated Three-Dose Regimens.**

| Dosing regimen | | 95% PI Lower Limit (days post-Dose 1) |
|---|---|---|
| Two-dose regimen:<br>Dose 1 – 480 mg piperaquine on Day 0<br>Dose 2 – 960 mg piperaquine 2 days post-Dose 1 | | 13.25 |
| Three-dose regimen:<br>Standard two-dose regimen, plus<br>3rd Dose (dose/days post-Dose 1) | | |
| 480 mg | Day 3 | 17.75 |
| | Day 4 | 18 |
| | Day 5 | 18.25 |
| | Day 6 | 18.75 |
| | Day 7 | 19.25 |
| | Day 8 | 19.5 |
| | Day 9 | 20 |
| 640 mg | Day 3 | 18.5 |
| | Day 4 | 18.75 |
| | Day 5 | 19.25 |
| | Day 6 | 19.5 |
| | Day 7 | 20 |
| | Day 8 | 20.75 |
| | Day 9 | 21.25 |
| 960 mg | Day 3 | 20.75 |
| | Day 4 | 21.25 |
| | Day 5 | 21.75 |
| | Day 6 | 22.25 |
| | Day 7 | 23 |
| | Day 8 | 23.5 |
| | Day 9 | 24 |

**Supplementary Table S3. Adverse Events by System Organ Class and Preferred Term**

| | **Primaquine Group (N=2)** | | | **Control Group (N=2)** | | | **Total (N=4)** | | |
|---|---|---|---|---|---|---|---|---|---|
| **System Organ Class<br>Preferred Term** | **n** | **(%)** | **E** | **n** | **(%)** | **E** | **n** | **(%)** | **E** |
| Subjects with at least one AE | 2 | (100) | 25 | 2 | (100) | 28 | 4 | (100) | 53 |
| **Blood and lymphatic system disorders** | **1** | **(50)** | **1** | **0** | **(0)** | **0** | **1** | **(25)** | **1** |
| Lymphadenopathy | 1 | (50) | 1 | 0 | (0) | 0 | 1 | (25) | 1 |
| **Gastrointestinal disorders** | **0** | **(0)** | **0** | **1** | **(50)** | **1** | **1** | **(25)** | **1** |
| Diarrhoea | 0 | (0) | 0 | 1 | (50) | 1 | 1 | (25) | 1 |
| **General disorders and administration site conditions** | **2** | **(100)** | **9** | **2** | **(100)** | **6** | **4** | **(100)** | **15** |
| Chills | 1 | (50) | 3 | 1 | (50) | 3 | 2 | (50) | 6 |
| Fatigue | 1 | (50) | 1 | 2 | (100) | 2 | 3 | (75) | 3 |
| Malaise | 1 | (50) | 3 | 0 | (0) | 0 | 1 | (25) | 3 |
| Pyrexia | 2 | (100) | 2 | 1 | (50) | 1 | 3 | (75) | 3 |
| **Infections and infestations** | **1** | **(50)** | **1** | **1** | **(50)** | **1** | **2** | **(50)** | **2** |
| COVID-19 | 1 | (50) | 1 | 0 | (0) | 0 | 1 | (25) | 1 |
| Rhinovirus infection | 0 | (0) | 0 | 1 | (50) | 1 | 1 | (25) | 1 |
| **Investigations** | **2** | **(100)** | **3** | **2** | **(100)** | **5** | **4** | **(100)** | **8** |
| Cardiac murmur | 1 | (50) | 1 | 0 | (0) | 0 | 1 | (25) | 1 |
| Haemoglobin decreased | 0 | (0) | 0 | 2 | (100) | 2 | 2 | (50) | 2 |
| Lymphocyte count decreased^ | 2 | (100) | 2 | 1 | (50) | 1 | 3 | (75) | 3 |
| Neutrophil count decreased | 0 | (0) | 0 | 1 | (50) | 1 | 1 | (25) | 1 |

| | Primaquine Group (N=2) | | | Control Group (N=2) | | | Total (N=4) | | |
|---|---|---|---|---|---|---|---|---|---|
| White blood cell count decreased | 0 | (0) | 0 | 1 | (50) | 1 | 1 | (25) | 1 |
| **Musculoskeletal and connective tissue disorders** | **2** | **(100)** | **3** | **1** | **(50)** | **1** | **3** | **(75)** | **4** |
| Arthralgia | 1 | (50) | 1 | 0 | (0) | 0 | 1 | (25) | 1 |
| Myalgia | 1 | (50) | 1 | 1 | (50) | 1 | 2 | (50) | 2 |
| Neck pain | 1 | (50) | 1 | 0 | (0) | 0 | 1 | (25) | 1 |
| **Nervous system disorders** | **2** | **(100)** | **4** | **1** | **(50)** | **7** | **3** | **(75)** | **11** |
| Dizziness | 2 | (100) | 2 | 0 | (0) | 0 | 2 | (50) | 2 |
| Headache | 1 | (50) | 2 | 1 | (50) | 7 | 2 | (50) | 9 |
| **Reproductive system and breast disorders** | **0** | **(0)** | **0** | **1** | **(50)** | **2** | **1** | **(25)** | **2** |
| Intermenstrual bleeding | 0 | (0) | 0 | 1 | (50) | 1 | 1 | (25) | 1 |
| Menstruation irregular | 0 | (0) | 0 | 1 | (50) | 1 | 1 | (25) | 1 |
| **Respiratory, thoracic and mediastinal disorders** | **2** | **(100)** | **2** | **1** | **(50)** | **2** | **3** | **(75)** | **4** |
| Cough | 2 | (100) | 2 | 0 | (0) | 0 | 2 | (50) | 2 |
| Oropharyngeal pain | 0 | (0) | 0 | 1 | (50) | 1 | 1 | (25) | 1 |
| Wheezing | 0 | (0) | 0 | 1 | (50) | 1 | 1 | (25) | 1 |
| **Skin and subcutaneous tissue disorders** | **0** | **(0)** | **0** | **1** | **(50)** | **3** | **1** | **(25)** | **3** |
| Hyperhidrosis | 0 | (0) | 0 | 1 | (50) | 3 | 1 | (25) | 3 |
| **Vascular disorders** | **1** | **(50)** | **2** | **0** | **(0)** | **0** | **1** | **(25)** | **2** |
| Hot flush | 1 | (50) | 2 | 0 | (0) | 0 | 1 | (25) | 2 |

^Decreases in lymphocyte count were rated as severe AEs (grade 3) in two participants: R002 on day 8 (0.45 x $10^9$/L) and R004 on day 10 (0.43 x $10^9$/L).
Normal range (1–2 x $10^9$ lymphocytes/L).
